# Enrichment of Rare Exonic Variants in Cognitive Superagers from Centenarian Families

**DOI:** 10.64898/2026.11.10.26365445

**Authors:** Lucas Fishbein, Kristina Mullin, Harold Bae, Isabel Castanho, Pourya Naderi, Vonetta M. Dotson, Winston Hide, Doo Yeon Kim, Paola Sebastiani, Susan Bookheimer, Stacy L. Andersen, Thomas T. Perls, Rudolph E. Tanzi, Dmitry Prokopenko, the Resilience/Resistance to Alzheimer’s Disease in Centenarians and Offspring (RADCO) Study

## Abstract

While the risk of cognitive impairment and dementia increases with advanced age, a fraction of individuals maintains high levels of cognitive function at extreme ages. Here, we compared whole-exome sequencing variant frequency data for 136 centenarian and offspring cognitive superagers, whose episodic memory performance matches normative expectations for persons ∼30 years younger, 114 average-cognition controls, and the gnomAD population reference database. A rare variant allele frequency analysis identified 114 rare coding variants significantly enriched in cognitive superagers, of which the ten most significant variants were exceedingly rare missense variants residing in nine genes implicating neurovascular biology (*LOX, CLEC4M, ZNG1F*), immune regulation (*HLA-B, HLA-DRB1, ITGAX*, *MUC4, MUC1*), and neuronal structure (*CEP131, two variants*). Together, these findings support the study of an extreme phenotype as a tractable framework for prioritizing rare coding variation and nominating a set of candidate loci for functional follow-up to help elucidate protective genetic mechanisms in cognitive superagers.

## Introduction

Advances in public health and medicine have markedly extended human life expectancy, illustrated by a 50% increase in the United States centenarian population from 2010 to 2020 ^1^ with projections estimating 3.2 million centenarians worldwide by 2050 ^2^. However, older age is the strongest risk factor for cognitive impairment, late-onset Alzheimer’s disease (AD) and vascular dementia. Alzheimer’s disease and related dementias (ADRD) risk rises steeply with age: prevalence in the US increases from 3% at ages 65–69 to 35% at 90 and older ^3^, reaches 53% in women and 45.5% in men beyond age 95 ^4^, and incidence climbs from 21.2% per year at ages 95–99 to 40.7% per year after 100 ^5^.

AD is a progressive neurodegenerative disorder characterized by profound cognitive impairment. Although anti-amyloid immunotherapies have recently led to modest slowing of clinical decline in early disease, treatment options remain limited ^6,7^. Early-onset AD (<65 years) is largely attributable to rare, highly penetrant variants ^8^. Late-onset AD is substantially heritable, with estimates of 58–79%, yet, beyond *APOE*, the strongest AD risk factor, common-variant genome-wide association studies (GWAS) identified roughly 150 AD-associated loci, which explain only about one-third of the inheritability of AD ^9–14^. Since standard GWAS pipelines typically filter out extremely rare variants, whole-exome sequencing has become a key approach for directly capturing rare protein-coding variation, implicating genes such as *TREM2, SORL1, ABCA7, ATP8B4, and ABCA1* ^15,16^.

Notably, reaching extreme old age is also highly heritable with recent heritability estimates of human life span over 50% ^17,18^. There are likely many different genetic signatures associated with survival to 100 that vary according to ancestry and prevalent non-genetic influences of exceptional longevity. Survival to even more extreme ages involves a more distinct and rare genetic profile ^19^, including enrichment of protective alleles and depletion of Alzheimer’s disease (AD) risk alleles, such as *APOE* ε4 ^19–22^.

Although the prevalence of cognitive decline and dementia is high in the older adult population, these conditions are not inevitable consequences of aging, as evidenced by the portion of the population that reaches age 100 and beyond, while maintaining relatively high levels of cognitive and physical function ^23,24^. These exceptional individuals raise fundamental questions about the biological mechanisms that preserve cognitive ability and delay dementia onset at very advanced ages, and whether genetics influence these protective features. Protective mechanisms may confer resistance to amyloid and tau accumulation in the brain or resilience to cognitive decline despite the presence of AD pathology ^25,26^. Because familial clustering of extreme longevity points to rare variants with larger effects than the modest common-variant associations, comparing extreme phenotypes has proven a powerful discovery strategy ^27–30^. For example, using cognitively healthy centenarians as controls nearly doubled the effect sizes of known AD variants ^22,31^, enabled a polygenic score for protection against AD ^32^, and, in a comparison of the top 1% of survivors with population controls, identified multiple longevity loci ^33^.

In the New England Centenarian Study (NECS), 12–25% of centenarians exhibit no symptoms of clinical dementia, and roughly 90% of those who eventually develop cognitive impairment remained cognitively intact well into their 90s ^24^. These individuals are not merely surviving longer; evidence from Amsterdam’s 100+ study shows they maintain prolonged periods of stable, high cognitive function ^34^ and are substantially more likely than cognitively impaired centenarians to survive to ages 105 and beyond ^35^.

An even smaller subset of centenarians, approximately 10% of the NECS centenarians, not only avoid cognitive decline and clinical dementia at advanced ages but have exceptional episodic memory, performing similarly to people 2-3 decades younger. Such individuals are called “cognitive superagers” ^36–38^. A substantial portion of their offspring likewise are “cognitive superagers” and exhibit approximately half the rate of age-related diseases and disability compared to their birth cohort ^35,39^, representing a related and materially informative opportunity for discovery ^40^. Numerous studies support the hypothesis that the exceptional cognitive preservation observed in extremely elderly individuals reflects active protective mechanisms that confer resistance or resilience to cognitive impairment and dementia at least through the mid-nineties ^17,41,42^, rather than reflecting the relative absence of neurodegenerative risk factors.

Here, we analyzed whole exome sequencing data from the Resilience/Resistance to Alzheimer’s Disease in Centenarians and Offspring (RADCO) study to identify protective genetic signals that support late-age cognitive preservation. By comparing rare variant allele frequencies across cognitive superagers, selected from centenarians and their offspring, whose episodic memory performance matches normative expectations for persons ∼30 years younger, with average-cognition RADCO participants as well as a population-level reference panel (gnomAD), we identified rare functional coding variants that are significantly enriched in cognitive superagers. These loci should help elucidate protective genetic mechanisms and associated biological pathways contributing to healthy brain aging.

## Results

The RADCO study is designed to identify the biological basis of cognitive resilience and resistance to AD in a cohort of centenarians, their offspring, and offspring spouses. RADCO participants are identified from ongoing studies of centenarians and their offspring (UH3AG064704: The Integrative Longevity Omics Study and U19AG023122: the Centenarian Project of the Longevity Consortium) and are classified into cognitive superagers, individuals whose performance on memory tasks matches normative expectations for persons ∼30 years younger, and individuals with average-cognition. A total of 256 RADCO participants (138 cognitive superagers and 118 subjects with average-cognition) met eligibility criteria to be included in the primary cognitive superager status endophenotype cohort, with 55 subjects being centenarians. After quality control and covariate exclusions (see Methods), 231 participants (129 cognitive superagers and 102 controls) were included for downstream single variant and gene-burden association analyses. The primary allele-frequency analysis included 250 RADCO participants (136 cognitive superagers and 114 controls) of European ancestry. In addition, we had access to a secondary endophenotype, cognitive status, available for 1292 WES subjects from the American Federation for Aging Research (AFAR) cohort. Demographic characteristics for both endophenotypes are summarized in Table 1.

**Table 1:** Demographic and inferred genetic ancestry information. **A.** RADCO participants by cognitive superager status, cases (n=138) and controls (n=118); **B.** 1,292 centenarians and offspring for which WES data was available from AFAR characterized by secondary endophenotype of cognitive status as cognitively normal (n=900) or cognitively impaired (n=392). The cognitive superager status as well as the cognitive status phenotype are defined in the methods section. Inferred genetic ancestry was assigned from WES-based optimal ancestry projection (OADP). Abbreviations: RADCO, Resilience/Resistance to Alzheimer’s Disease in Centenarians and Offspring; AFAR, American Federation for Aging Research, AFR, African; AMR, Admixed American; EAS, East Asian; EUR, European; SAS, South Asian; SD, standard deviation.

| Phenotype | Group | n (centenarian n, %) | Age, mean (SD), years | Gender, n (%) | Inferred genetic ancestry (n) |
|---|---|---|---|---|---|
| **A. RADCO Cognitive superager status** | Cognitive superagers (cases) | 138 (20, 14.5%) | 78.92 (11.61) | Female: 112 (81.2%)<br>Male: 26 (18.8%) | EUR: 136, AFR: 2, AMR: 0, EAS: 0, SAS: 0 |
|  | Average-cognition (controls) | 118 (35, 29.7%) | 86.84 (13.11) | Female: 61 (51.7%)<br>Male: 57 (48.3%) | EUR: 114, AFR: 2, AMR: 1, EAS: 1, SAS: 0 |
|  | All subjects with superager status | 256 (55, 21.5%) | 82.43 (12.89) | Female: 173 (67.6%)<br>Male: 83 (32.4%) | EUR: 250, AFR: 4, AMR: 1, EAS: 1, SAS: 0 |
| **B. AFAR Cognitive status** | Cognitively normal (cases) | 900 (220, 24.4%) | 86.03 (12.38) | Female: 574 (63.8%)<br>Male: 326 (36.2%) | EUR: 867, AFR: 19, AMR: 14, EAS: 0, SAS: 0 |
|  | Cognitively impaired (controls) | 392 (287, 73.2%) | 99.67 (8.28) | Female: 293 (74.7%)<br>Male: 98 (25.0%)<br>Unknown: 1 (0.3%) | EUR: 369, AFR: 15, AMR: 6, EAS: 1, SAS: 1 |
|  | All subjects with cognition status | 1292 (507, 39.2%) | 90.17 (12.92) | Female: 867 (67.1%)<br>Male: 424 (32.8%)<br>Unknown: 1 (0.08%) | EUR: 1,236, AFR: 34, AMR: 20, EAS: 1, SAS: 1 |

We first carried out a single-variant association analysis for the binary cognitive superager endophenotype using REGENIE ^43^. After variant-level QC, 418,278 variants were analyzed with a minimum minor allele count (MAC) of 5. Across the exome-wide scan, 8,690 tests required Firth correction, no tests failed, and 254,895 variants were excluded for low MAC, leaving 163,383 effective tests. No variant reached the Bonferroni-corrected threshold of P = 3.06 × 10^−7^ (Figure 1A), most likely due to the modestly-sized discovery cohort of only 231 individuals. Top variants included four *DBH* missense variants in LD with each other, with P-values ranging from 2.50 × 10^−6^ to 1.50 × 10^−5^, with b between –2.76 and –2.52 (depleted in cognitive superagers). Other top variants include four *PATJ* variants (P = 4.10 × 10^-5^ to 4.85 × 10^−4^, b = -0.96 to -0.85), *SPATA31A1* (P = 4.10 × 10^−5^, b = +1.02), *HLA-DOB* (P = 5.90 × 10^-5^, b = –2.57), *SLC25A23* (P = 6.04 × 10^-5^, b = +0.90), and two *MAP3K10* variants (P = 6.25 × 10^-5^ and 1.49 × 10^-4^, b = +0.98 and +0.91). A full breakdown of the top 40 single-variant hits ranked by P-value is provided in Supplementary Table B1.

**Figure 1:**
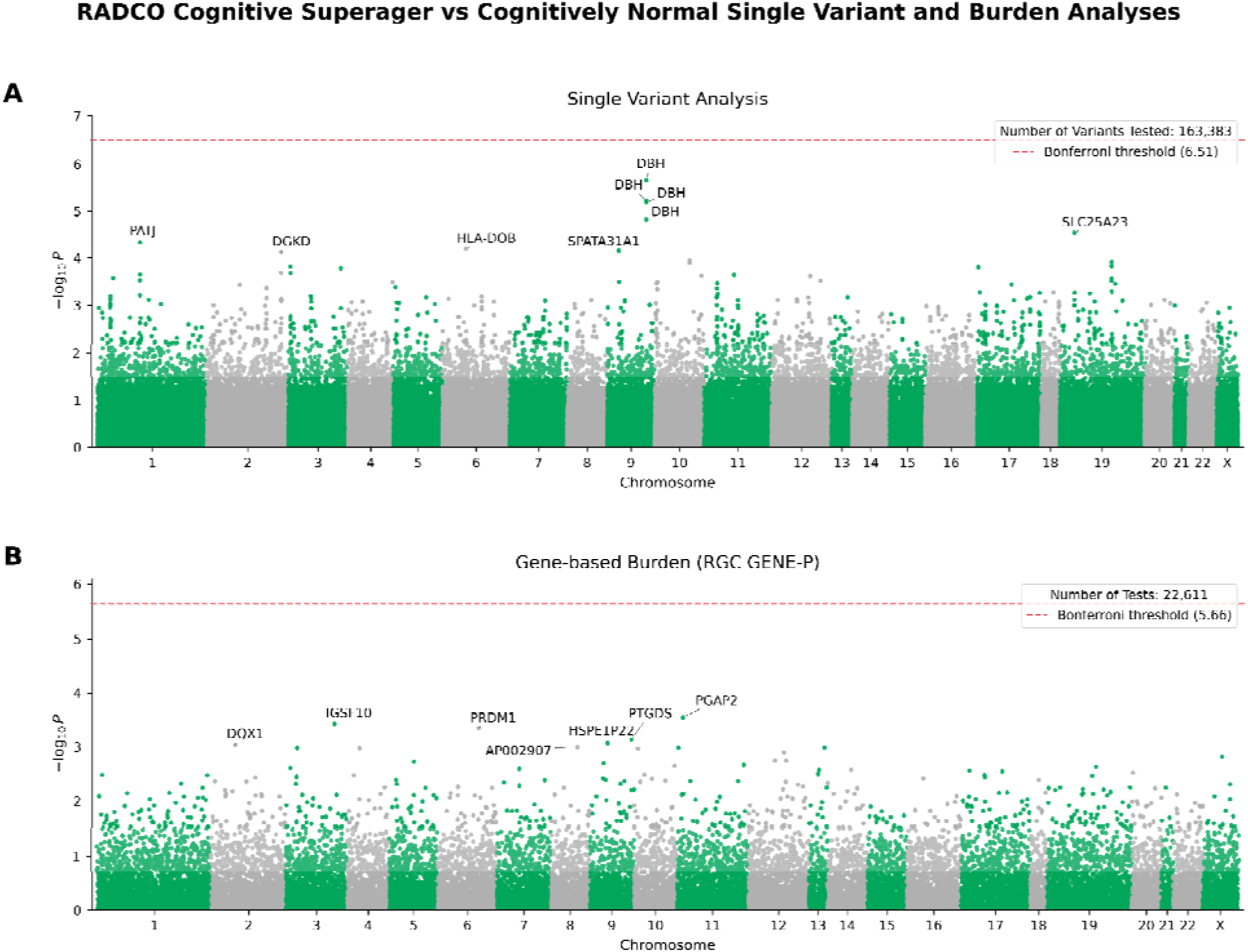
RADCO cognitive superager vs average cognitive performance exome-wide single-variant and gene-based burden association analyses. REGENIE association results comparing RADCO cognitive superagers with RADCO average-cognition individuals (n = 231). (A) Single-variant Manhattan plot showing –log_10_(P) across exome variants passing minor-allele-count (MAC) filters; the analysis had a genomic control inflation factor of λ = 1.07. The dashed red line marks the Bonferroni significance threshold (P = 3.06×10^−7^; –log_10_(P) = 6.51). (B) Gene-level RGC GENE-P burden Manhattan plot across 22,611 gene sets; genomic control inflation factor λ = 1.02. The dashed red line marks the Bonferroni significance threshold (P = 2.21×10^−6^; –log_10_(P) = 5.66). No gene exceeded Bonferroni correction.

Next, we performed a gene-based rare-variant burden analysis of our primary binary cognitive superager endophenotype. The analysis aggregated 459,705 variants into 29,376 gene sets. We report REGENIE’s RGC-GENE-P omnibus statistic, which combines burden-style and variance-component signals (ACATV and SKATO-ACAT) via ACAT. RGC-GENE-P produced results for 22,611 of the gene sets, across the genome-wide run, 6,131 tests used REGENIE’s approximate Firth stabilization, no tests failed, and no tests were excluded for low MAC. Using a Bonferroni correction across 22,611 gene-level GENE_P tests (α = 0.05 / 22,611 = 2.21 × 10^−6^; −log_10_(p) = 5.66), no gene exceeded the exome-wide threshold (Figure 1B). Several genes showed suggestive gene-level associations (P < 1 × 10^−3^). The strongest signals were *PGAP2*, *IGSF10, PRDM1, PTGDS* (P ranging from 2.84 × 10^−4^ to 7.19 × 10^−4^). For each of these loci, the strongest contributing signal came from the OtherMask annotation stratum. Supplementary Table B3 lists the top gene sets ranked by GENE_P −log_10_(P). Among 150 loci identified in AD GWAS meta-analyses ^9,10,13^, *DOC2A* showed nominal single-variant association (P = 0.0345, rs1140239), while *EPHA1* and *NCK2* showed nominal burden associations (P = 0.00411 and 0.0437; Supplementary Tables B1 and B3).

We next searched for enrichment of extremely rare exonic variants in cognitive superagers. For this purpose, we focused on rare variants that had a minor allele frequency less than 0.001 in the non-Finnish European (NFE) subset of gnomAD v4.1 ( AF_NFE_ < 0.001) and compared allele frequency in 136 European cognitive superagers versus 114 European average-cognition controls as well as the gnomAD v4.1 NFE reference group. Beginning with 1,224,073 whole-exome-captured variants screened across RADCO and gnomAD NFE, we used Fisher’s exact test to compare enrichment in cognitive superagers (Methods).

We first identified 976 rare variants that were carried by ≥4 cognitive superagers and were significantly enriched in cognitive superagers as compared to gnomAD NFE (P_adj_ < 0.01) but not enriched in the RADCO average-cognition cohort compared with gnomAD NFE (P_adj_ ≥ 0.01). For the direct comparison of RADCO cognitive superagers with RADCO average-cognition participants, no variant remained significant after Benjamini–Hochberg correction across 870,018 rare variants (all P_adj_ = 1; smallest unadjusted P = 4.29 × 10^−6^). Given the limited number of rare alleles observed in 250 individuals, this comparison was underpowered and was not used to prioritize variants. The resulting distribution of these rare variants reflected the exome-capture footprint (Figure 2). The candidate missense-to-synonymous ratio (1.45:1) did not significantly differ from that of the 870,018 frequency-matched rare exonic variants in the same cohort (1.59:1; OR = 0.91, Fisher’s exact P = 0.43), indicating that the functional-class composition of the candidate set is typical of rare exonic variation; full details on this analysis are in Supplementary Table B4.

**Figure 2:**
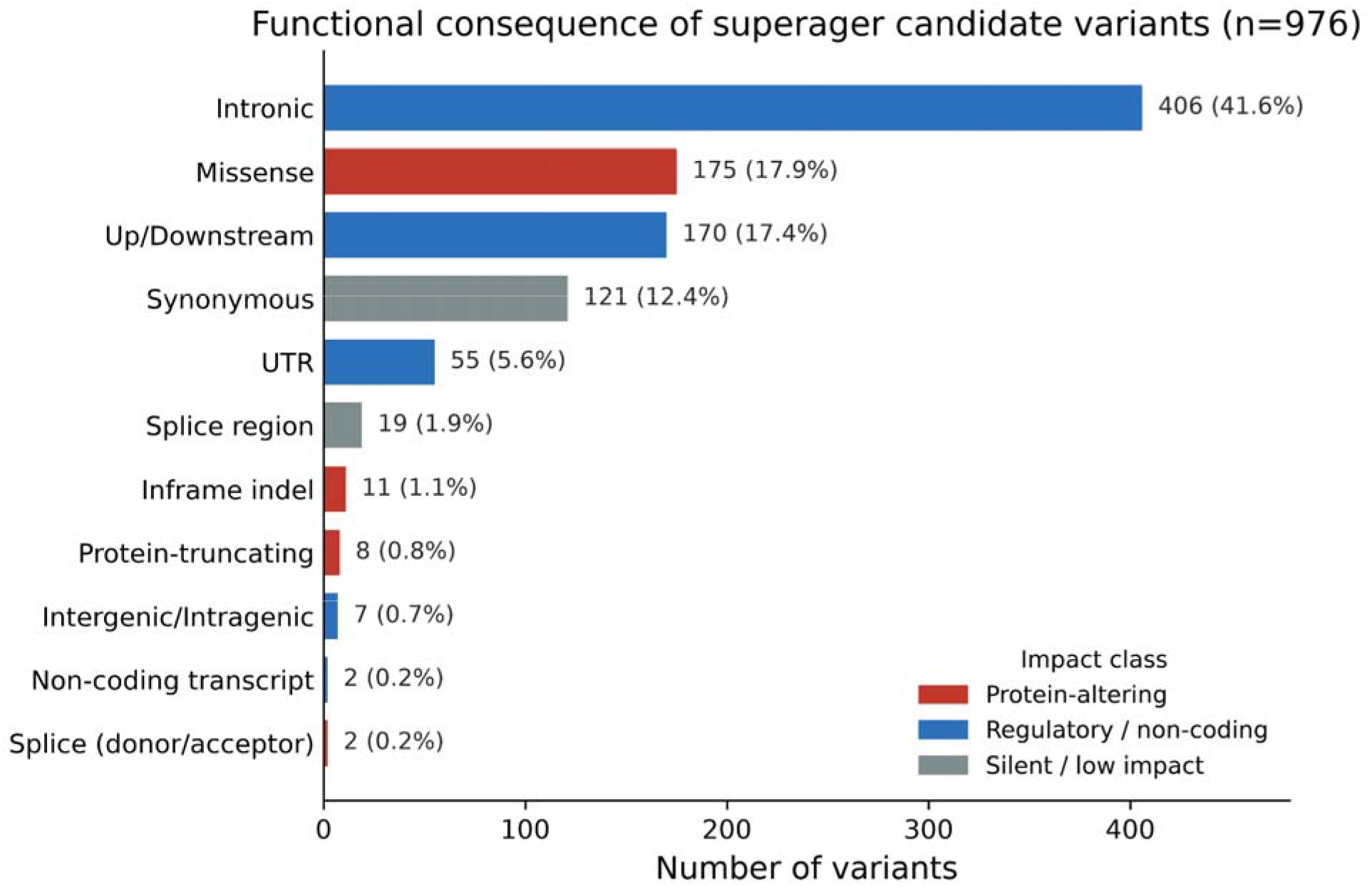
Functional-consequence distribution of the 976 cognitive superager candidate variants. Rare exonic variants (gnomAD AF_NFE_ < 0.001) carried by ≥4 cognitive superagers that met minor-allele-frequency criteria annotated with SnpEff and grouped into higher-level consequence classes. Bars show the number of variants per class (with percentage of the 976 total) and colored by predicted impact tier: protein-altering (protein-truncating, splice donor/acceptor, inframe indel, missense; red), silent/low-impact (synonymous, splice region; grey), and regulatory/non-coding (intronic, up/downstream, UTR, intergenic, non-coding transcript; blue). AF, allele frequency; NFE, non-Finnish European; UTR, untranslated region.

Restricting candidates to moderate-to-high predicted impact and excluding single-lineage or Hardy-Weinberg-deviating variants and completing our variant filtering pipeline as described in Figure 3 yielded 114 significant candidates, full details in Supplementary Table B5. We stratified the 114 variants by the stringency of the cognitive-superager-vs-NFE enrichment into tier 1 (P_adj_ < 1×10^−8^ versus gnomAD NFE; n = 10) and tier 2 (1×10^−8^ ≤ P_adj_ < 0.01 versus gnomAD NFE; n = 104), with tier 1 representing the most conservatively enriched candidates. The 114 variants mapped to 104 protein-coding genes, and 42 (37%) were absent from RADCO average-cognition controls. Carrier-overlap analysis indicated shared signals at *CEP131*, *CFH*, *KRTAP10-5*, and *HLA-DRB1* but independent signals at *MUC4* and *KRTAP10-6*. All ten tier 1 variants were missense mutations and five had an alternative allele count of zero in the RADCO average-cognition cohort (Table 2).

**Figure 3:**
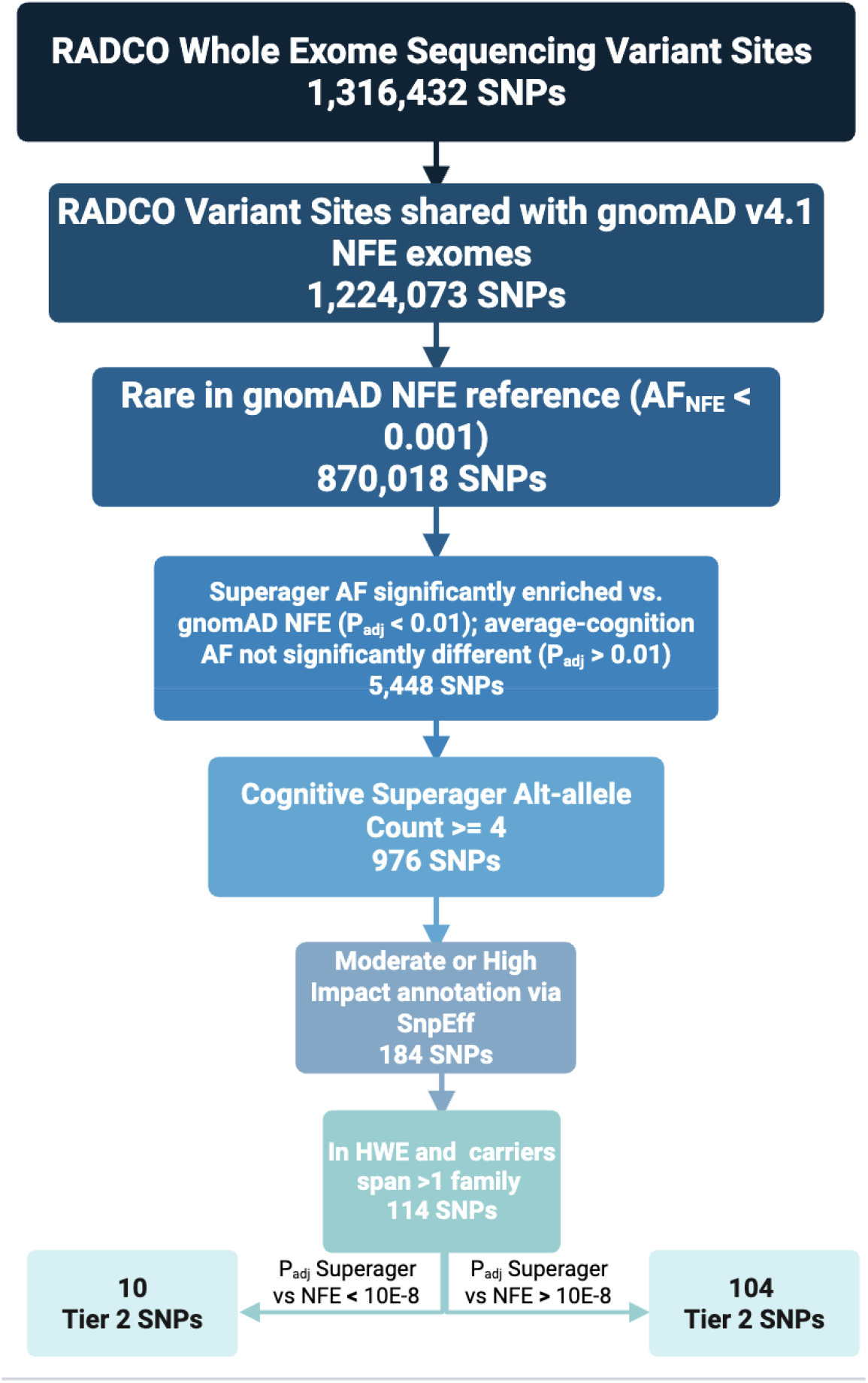
Variant filtering workflow for RADCO superager allele-frequency analysis. Starting from whole-exome sequencing (WES) variant sites in the RADCO cohort (1,316,432 SNPs) and then merging with gnomAD exome data, variants were retained if they passed population screening across all study groups (1,224,073 SNPs), then restricted to rare variants (870,018 SNPs). Sites were kept if superager allele frequency differed from gnomAD non-Finnish European (NFE) reference frequencies while the cohort average-cognition allele frequency did not (5,448 SNPs). Additional filters required a cognitive superager minor-allele count ≥4 (976 SNPs), protein-altering consequence (184 SNPs), not deviating from Hardy–Weinberg equilibrium (HWE) in the study sample at a threshold of 1 x 10^-6^, and representation in more than a single-family lineage (114 SNPs). The final set was stratified by adjusted P-value for superager vs. NFE comparison: tier 1, P_adj_ < 1×10^−8^ (n = 10 SNPs); tier 2, P_adj_ ≥ 1×10^−8^ (n = 104 SNPs). AF, allele frequency; HWE, Hardy–Weinberg equilibrium; NFE, non-Finnish European; SNP, single-nucleotide polymorphism; WES, whole-exome sequencing.

**Table 2:** Tier 1 rare candidate variants from RADCO cognitive superager rare-variant allele-frequency analysis. Tier 1 variants are those passing the more stringent significance threshold of P_adj_ < 1 × 10^−8^ in the Cognitive-Superager-vs-gnomAD-NFE comparison while showing non-significantly different allele frequencies between the RADCO average-cognition control group and the NFE reference (P_adj_ > 0.01; see Methods). Abbreviations: AC, allele count (number of alternative alleles observed); AN, allele number (total observed alleles at the locus); AF, alternative allele frequency; NFE, Non-Finnish European; P_adj_, multiple-testing-corrected Fisher’s exact P-value.

|  |  |  | Fisher’s exact P<sub>adj</sub> |  | Alternative allele frequency (Allele Count / Alleles observed) |  |  |
|---|---|---|---|---|---|---|---|
| Locus | RsID | Variant | Cognitive superager vs NFE | Average-cognition vs NFE | gnomAD NFE | RADCO cognitive superager | RADCO average-cognition |
| *ITGAX* | rs2142487206 | A242V | 2.78 × 10<sup>−16</sup> | 1.90 × 10<sup>−2</sup> | 9.01 × 10<sup>−7</sup><br>(1 / 1,109,692) | 2.66 × 10<sup>−2</sup><br>(5 / 188) | 6.67 × 10<sup>−3</sup><br>(1 / 150) |
| *ZNG1F* | rs66626885 | L220I | 5.68 × 10<sup>−13</sup> | 7.78 × 10<sup>−2</sup> | 1.09 × 10<sup>−5</sup><br>(12 / 1,104,748) | 2.23 × 10<sup>−2</sup><br>(5 / 224) | 5.32 × 10<sup>−3</sup><br>(1 / 188) |
| *MUC1* | rs1667657598 | P423T | 1.98 × 10<sup>−12</sup> | 6.36 × 10<sup>−2</sup> | 4.97 × 10<sup>−6</sup><br>(1 / 201,390) | 2.36 × 10<sup>−2</sup><br>(5 / 212) | 6.10 × 10<sup>−3</sup><br>(1 / 164) |
| *CEP131* | rs141346984 | H238R | 4.91 × 10<sup>−12</sup> | 1 | 1.37 × 10<sup>−4</sup><br>(152 / 1,111,888) | 2.61 × 10<sup>−2</sup><br>(7 / 268) | 0<br>(0 / 222) |
| *HLA-B* | rs148138285 | D201Y | 1.01 × 10<sup>−11</sup> | 1 | 1.24 × 10<sup>−6</sup><br>(1 / 805,222) | 1.65 × 10<sup>−2</sup><br>(4 / 242) | 0<br>(0 / 204) |
| *MUC4* | rs1269489956 | V3971L | 3.01 × 10<sup>−11</sup> | 1 | 8.69 × 10<sup>−5</sup><br>(56 / 644,092) | 2.59 × 10<sup>−2</sup><br>(6 / 232) | 0<br>(0 / 182) |
| *CEP131* | rs143331394 | S642G | 1.04 × 10<sup>−10</sup> | 1 | 2.18 × 10<sup>−4</sup><br>(242 / 1,111,858) | 2.61 × 10<sup>−2</sup><br>(7 / 268) | 0<br>(0 / 222) |
| *LOX* | rs141394149 | S132F | 1.45 × 10<sup>−10</sup> | 3.71 × 10<sup>−1</sup> | 1.02 × 10<sup>−4</sup><br>(113 / 1,111,802) | 2.24 × 10<sup>−2</sup><br>(6 / 268) | 4.50 × 10<sup>−3</sup><br>(1 / 222) |
| *HLA-DRB1* | rs16822820 | S66P | 9.99 × 10<sup>−10</sup> | 1 | 1.05 × 10<sup>−5</sup><br>(10 / 950,496) | 1.60 × 10<sup>−2</sup><br>(4 / 250) | 0<br>(0 / 206) |
| *CLEC4M* | rs113080783 | Q118R | 1.54 × 10<sup>−9</sup> | 3.34 × 10<sup>−2</sup> | 1.62 × 10<sup>−4</sup><br>(170 / 1,048,258) | 2.34 × 10<sup>−2</sup><br>(6 / 256) | 9.09 × 10<sup>−3</sup><br>(2 / 220) |

The majority of the variants in the set were missense mutations (109/114), with five predicted protein-truncating variants (two frameshift, two stop-gained, and one stop-gained/splice-region). Exploratory over-representation analysis of the 114 genes did not identify a single dominant pathway but showed nominal convergence on immune and antigen-presentation processes (interferon-γ signaling; *HLA-B*/-*C*/-*DRB1*/-*DQB2*, *PTPN2*), mucin-type O-glycosylation (*MUC1/MUC4/MUC21*), and complement/phagosome function (*CFH*, *CFHR5*, *ITGAX*, *F5*), full details in Supplementary Table B8.

We next predicted functional genomic impact using AlphaGenome for all 114 rare variants (Supplementary Table B6). Tier 1 variants in *LOX* and *CLEC4M* showed predicted chromatin or expression effects, and immune-associated *HLA-B*, *HLA-DRB*1, and *ITGAX* variants showed predicted promoter or splice effects. Among tier 2 variants, loss-of-function variants in *TNFSF18* and *HMMR* had the strongest predicted splice disruption.

Because rare variants at *HLA-B* and *HLA-DRB1* were among the top candidates, we next asked whether classical HLA alleles at these loci are associated with cognitive health more broadly. For this we used the larger AFAR cohort, which had WGS imputed data available. Since cognitive superager status was not determined in this AFAR cohort, we utilized the secondary cognitive normal vs cognitively impaired endophenotype and imputed classical HLA alleles in 772 participants in the AFAR sample with complete covariate data, 280 of which were centenarians. This analysis is therefore complementary to, rather than a replication of, the rare-variant findings. Under additive coding, no lead allele met 7-locus FDR significance (all q ≥ 0.073; Supplementary Table B7). Nominal associations (P < 0.05) were observed for *HLA-DPB1*\*02:01 (OR = 0.64, P = 0.011), *HLA-DRB1*\*04:01 (OR = 1.57, P = 0.037), and *HLA-DQA1*\*01:03 (OR = 0.65, P = 0.038). In a dominant carrier sensitivity analysis, all nominally significant p-value signals remained with the addition of *HLA-C*\*07:01 showing nominal significance (OR 1.49, P = 0.032) in the opposite direction of our additive results. *HLA-DPB1*\*02:01 remained significant after 7-locus FDR correction (OR = 0.58, P = 0.006, q = 0.040; Supplementary Table B7). No additional independent signals were identified.

## Discussion

Here we present the largest whole-exome sequencing analyses of clinically defined cognitive superagers conducted to date. Using complementary single-variant, gene-based burden, and extreme-phenotype allele-frequency frameworks, we identified 114 rare coding variants significantly enriched in cognitive superagers relative to population-scale reference controls. We highlight the top ten tier 1 candidate variants, which converge on three biologically coherent themes: immune regulation, neurovascular biology, and neuronal structure.

While standard single variant-level and gene-based burden analyses did not identify exome-wide significant associations with the cognitive superager phenotype, likely due to low sample size, we observed suggestive signals at several loci. These include *DBH*, *SLC25A23, PATJ, HLA-DOB*, and *SPATA31A1* in the single-variant analysis, and *PGAP2, IGSF10, PRDM1, PTGDS, HSPE1P22, DQX1,* and *AP002907.1* in the gene-based burden analysis. *DBH* was the leading single-variant locus, with four linked variants depleted in cognitive superagers. *DBH* encodes dopamine beta-hydroxylase, which converts dopamine to norepinephrine. Previous studies have reported small effects of DBH on AD risk, with stronger interaction signals by sex, inflammatory context, and gene-gene epistasis ^44,45^, or by AD biomarkers such as CSF p-tau181 ^46^. The tight junction scaffolding gene *PATJ* has been linked to frailty phenotypes^47^ related to broader age-related physiology. *PGAP2* was the leading burden locus. Biallelic hypomorphic *PGAP2* variants disrupt GPI-anchor remodeling and cause neurodevelopmental syndromes, supporting neurological relevance but not an established role in AD or cognitive ^48,49^. Suggestive signals at the immune regulator PRDM1 and the prostaglandin D2 synthase PTGDS provide additional hypotheses for follow-up ^50–53^.

The strongest findings arose from the two-way allele-frequency analysis, which required enrichment in cognitive superagers versus gnomAD NFE while controls remained concordant with gnomAD NFE. After functional, family-lineage, and Hardy-Weinberg filters, 114 significant candidates were retained (10 conservative tier 1 and 104 broader tier 2). This design reduces cohort-wide technical or ancestry signals. An exploratory over-representation analysis of the 114 genes did not resolve a single dominant pathway; rather, it converged nominally on immune and antigen-presentation processes, mucin-type O-glycosylation, and complement/phagosome function, supporting the biological themes that dominate the tier 1 variants discussed below.

Our ten tier 1 variants span three broad functional themes: neurovascular biology, immune regulation, and neuronal structure. The theme of neurovascular biology is represented by *LOX* (vascular matrix crosslinking, CAA), *ZNG1F* (intracranial aneurysm expression signature), and *CLEC4M* (endothelial clearance). This theme is consistent with a large body of evidence linking preserved cerebrovascular integrity to cognitive aging ^54,55^.

*LOX* and its paralogs *LOXL1*–*LOXL4* are extracellular amine oxidases that catalyze collagen and elastin cross-linkage, thereby setting vessel-wall tissue stiffness and governing the matrix remodeling that maintains vascular integrity ^56,57^. Increased LOX-family signaling has been linked with senescence-associated phenotypes, including fibrosis and vascular remodeling ^58^. Directly relevant to the neurovascular theme, *LOX* has been reported to localize with amyloid-beta pathology and is associated with cerebral arteriolar stiffening and degeneration ^59–61^. Along these lines, pharmacological inhibition of *LOX* has been proposed as a strategy to improve intramural periarterial drainage and reduce the accumulation of Aβ in the parenchyma and within blood vessel walls ^62^. In silico profiling with AlphaGenome predicts a large histone-modification effect for the cognitive superager-enriched *LOX* variant, S132F, in brain and neurons, consistent with the possibility that this allele affects chromatin state at a locus already implicated in vascular matrix remodeling. S132F resides in the propeptide and replaces a small polar residue with a bulky hydrophobic one suggesting a possible effect on precursor handling versus mature-enzyme function.

*CLEC4M* encodes *CD209L*, a sinusoidal endothelial C-type lectin involved in vascular ligand handling and clearance at the immune-endothelial interface ^63^. Human genetic and functional studies have shown that *CLEC4M* regulates circulating coagulation factor levels ^64–66^. We found the rare *CLEC4M* variant, Q118R, enriched in cognitive superagers and showed moderate predicted RNA-seq and histone-modification scores in AlphaGenome, supporting continued evaluation of this locus as a vascular-immune interface candidate. Q118R adds a positive charge to the extracellular domain possibly affecting receptor oligomerization. *ZNG1F* was also identified among genes with elevated intraluminal expression in human intracranial aneurysms, nominating it as a tentative marker of cerebrovascular vessel-wall pathology ^67^. The enriched rare *ZNG1F* missense mutation, L220I, might possibly affect the metal-ion binding zinc-chaperone or GTPase activities of this protein.

With regard to the immune regulation theme, the HLA findings align with prior associations of this region with longevity and neurodegeneration. The nominal association of the classical allele *HLA-DRB1*\*04:01 with the secondary phenotype (cognitive status) was directionally concordant with its reported protective association with AD ^68^, while the FDR-significant *HLA-DPB1*\*02:01 dominant-model result may represent a novel cognitive-aging signal requiring replication. In addition, Joshi et al. ^20^ reported an association of *HLA-DQA1/DRB1* with longevity. Both HLA gene rare variants exhibit strong enrichment in cognitive superagers relative to gnomAD NFE and are absent in the RADCO average-cognition subjects. The enriched *HLA-B* missense mutation, D201Y, resides at the end of the extracellular α2 domain and may potentially alter local stability, peptide presentation, or receptor interactions. The S66P HLA-DRB1 missense mutation enriched in cognitive superagers is a non-conservative change in the peptide-binding β1 domain and may potentially affect antigen specificity.

*ITGAX* encodes the CD11c integrin alpha-X subunit, which pairs with CD18 to form complement receptor 4 (CR4), a defining marker of a microglial subpopulation that accumulates around amyloid plaques and has been characterized as the “disease-associated microglia” (DAM) signature in mouse and human AD brain ^69,70^. Although *ITGAX* has not been identified as an AD-risk locus in genome-wide association studies, the central role of CD11c-marked microglia in the response to amyloid pathology makes it a biologically plausible candidate for influencing late-life cognitive trajectory. The enriched *ITGAX* missense mutation, A242V, resides in the extracellular αI ligand-binding domain could potentially affect ligand affinity.

AlphaGenome predicted moderate promoter and chromatin effects for the tier 1 immune variants *HLA-B*, *HLA-DRB1*, and *ITGAX*. Among tier 2 candidates, the *TNFSF18* frameshift showed near-maximal predicted splice disruption (0.99), suggesting that some non–tier 1 hits may act through transcript instability or loss of function rather than missense effects alone. Functionally, these immune findings are aligned with a model in which HLA-mediated antigen presentation, complement-receptor engagement, and microglial activation jointly modulate maladaptive neuroinflammation, offering a plausible mechanism for preserved cognition in advanced age. Altered antigen presentation could influence microglial activation states, complement deposition on synapses, and clearance of amyloid-β, all of which are implicated in AD pathogenesis ^71,72^.

The mucin family is represented by two tier 1 variants in *MUC1* and *MUC4*. Both genes encode large transmembrane mucins best characterized at epithelial barriers and in cancer, organ-specific fibrosis, and chronic inflammation ^73–77^. *MUC1* domains have also been shown to physically interact with immune response-related proteins, including receptors for advanced glycation end products (RAGE), major histocompatibility complex II (MHC-II), and extracellular matrix receptor integrin alpha 2 (*ITGA2*). The cognitive superager-enriched *MUC1* missense mutation, P423T introduces a potential O-glycosylation acceptor. *MUC4* is a large cell-surface mucin involved in epithelial protection, adhesion-related behavior, and ERBB2-associated signaling. The enriched *MUC4* missense mutation, V3971L, might potentially affect Erb-B2 receptor tyrosine kinase 2 (ERBB2) signaling. Interestingly, ERBB2 (HER2) has been implicated in AD as a suppressor of autophagic clearance of APP C-terminal fragment C99, thereby enhancing Aβ production ^78^. *MUC1* and *MUC4* may also affect the function of Erb-B2 receptor tyrosine kinase 4 (ERBB4), which has recently been shown to exacerbate tau-tangle propagation in response to amyloid pathology in excitatory neurons ^79^. Thus, these two genes may be directly related to resilience to AD pathology.

The third functional theme, neuronal structure, is represented by *CEP131* (centrosome function, axonogenesis), in which two missense variants were found to be enriched in cognitive superagers. Both of these *CEP131* variants were consistently observed together in 7 subjects, suggesting strong linkage disequilibrium. *CEP131* has previously been implicated in frontotemporal dementia (FTD) with suggestive SNPs pertaining to both protective and risk haplotypes ^80^, implicating impaired neuronal development, maturation, and axonogenesis. *CEP131* has also been implicated in Parkinson’s disease pathogenesis via de novo mutations ^81^. Functionally, the missense mutation, S642G, increases backbone flexibility and could therefore affect protein interactions, or possibly eliminate a phosphorylation site. The other mutation, H238R, may potentially alter electrostatic interactions and binding to centrosomal partners.

A key strength of this study is the use of an extremely rare, robustly characterized phenotype centered on cognitive superagers, a subset of which have already reached extreme ages of 100+ years, in order to detect enrichment of extremely rare variants ^82–84^. This population is inherently enriched for protective genetic architecture, thus this design concentrates biological signals in a manner not achievable in standard case-control studies ^22^. Our pairwise Fisher’s tests, across three groups design, minimized possible sequencing artifacts from the RADCO dataset.

Our study is not without limitations. Although it represents one of the largest rare variant analyses of cognitive superagers conducted to date, statistical power is constrained by the modest cohort size. Accordingly, the direct comparison within RADCO of cognitive superagers with average-cognition controls identified no variants after multiple-testing correction (smallest unadjusted P = 4.29 × 10^−6^), so evidence for enrichment rests on the comparison with the external gnomAD reference, which is subject to cohort-specific technical differences. It is important to note that Fisher’s exact tests cannot be adjusted for covariates. However, the extreme-phenotype enrichment framework was chosen because the Fisher’s-exact framework retains full statistical resolution under sparse 2 x 2 tables ^85,86^. It is also important to note that due to the high inter-sample size imbalance when comparing the gnomAD NFE reference population (n = ∼500,000) to our cognitive superager subgroup (n = 136), odds ratios from Fisher’s exact tests may be unstable. Finally, to minimize confounding from population stratification, we restricted the current analysis to unrelated individuals of predominantly European ancestry. Expanding future efforts to more ancestrally diverse populations will be critical to detect population-specific variation and ensure that insights into genetically based optimal cognitive aging are broadly generalizable across human populations.

Collectively, our findings support a model in which cognitive superaging reflects active genetic protection rather than a mere absence of risk. More broadly, our results demonstrate that extreme-phenotype sampling is a tractable and biologically informative strategy for accessing protective genetic architecture and establishing a focused roadmap for the mechanistic dissection of cognitive resilience.

## Methods

### Study Populations

The Resilience/Resistance to Alzheimer’s Disease in Centenarians and Offspring (RADCO) eligible participants are identified from ongoing studies of centenarians and their offspring (UH3AG064704: The Integrative Longevity Omics Study and U19AG023122: the Centenarian Project of the Longevity Consortium). RADCO seeks to determine the protective factors and underlying biological mechanisms that confer this extreme phenotype of optimal cognitive aging. The RADCO sample is comprised of centenarian cognitive superagers, centenarians with average-cognition, centenarian offspring cognitive superagers, offspring with average-cognition, and spouses of the offspring as a referent cohort. Phenotypic measures include sociodemographic, medical history, and lifestyle questionnaires; a neuropsychological assessment; blood pressure and grip strength measures; accelerometry; biospecimen collection (i.e., blood and stool); and an informant interview.

Participants in RADCO are classified as “cognitive superager” or “average-cognition” based on neuropsychological test performance criteria. Cognitive superagers performed at or above the normative mean for someone 30 years younger on the Hopkins Verbal Learning Test-Revised (HVLT-R) delayed recall, and at least average performance relative to same-aged peers on non-memory tests (i.e., semantic fluency [animals] and letter fluency [FAS]. The HVLT-R delayed recall cutoffs were ≥ 10 words for offspring and spouses and ≥ 8 words for centenarians. Participants with average-cognition performed within the average range (i.e., 1 standard deviation for offspring and 1.5 standard deviations for centenarians) on HVLT-R delayed recall and the non-memory tests compared to same-aged peers. Because episodic memory normative data are not available for centenarians, average values for nonagenarians were used instead.

This primary endophenotype consists of 256 subjects from whom whole exome sequence data were generated. 138 were cognitive superagers and 118 subjects classified as having average-cognition based on the above definitions. This sample includes 55 centenarians, of which 20 were cognitive superagers and 35 had average-cognition (Table 1A).

Downstream association analyses used WES data from the American Federation for Aging Research (AFAR) sample of centenarians and offspring and a secondary binary endophenotype of cognitive status (cognitively normal vs cognitively impaired) derived from standardized cognitive screener and/or informant interviews (i.e., a Dementia Questionnaire) ^87^. The secondary cognitive-status analysis included 507 centenarians (39.2% of 1,292 participants), comprising 220 cognitively normal participants (MMSE ≥ 24) and 287 cognitively impaired (MMSE ≤ 23) (Table 1B).

Cognitive status was determined by one or more of the following: the Mini-Mental State Examination ^88^, the Telephone Interview for Cognitive Status - Modified ^89,90^, and the Blessed Information–Memory–Concentration Test ^91^. Established cut-points were used to dichotomize each instrument: MMSE ≤ 23 was coded as impaired and ≥ 24 as normal; TICS-M ≤ 25 as impaired and ≥ 26 as normal; BIMC ≤ 26 as impaired and ≥ 27 as normal ^92^; and/or endorsement of memory problems and functional impairments on the Dementia Questionnaire according to the algorithm by Kawas et al. ^93^. Assessments marked as participant refusal or invalid due to hearing, environmental, or dispositional factors were excluded. Administrations coded as “unable due to dementia” were coded as impaired. The resulting binary cognitive-status variable was used as the secondary endophenotype in downstream association analyses, yielding 900 cognitively normal and 392 cognitively impaired participants.

### Study Data and Pre-Processing

Whole Exome Sequencing (WES) was performed via funding from the American Federation for Aging Research (AFAR) using samples from multiple centenarian and offspring studies. Briefly, exome capture was performed using a custom reagent kit from Kapa Biosystems using a fully automated approach at Regeneron. A unique 6-base pair barcode was added to each DNA fragment during library preparation to facilitate multiplexed exome capture and sequencing. Multiplexed samples were sequenced using 75 bp paired-end sequencing on an Illumina v4 HiSeq 2500 to a coverage depth sufficient to provide greater than 20x haploid read depth. Raw sequence data from each Illumina HiSeq 2500 run were uploaded to DNAnexus for sequence read alignment and variant identification. Raw sequence data were converted from BCL files to sample-specific FASTQ-files, which were aligned to human reference build GRCh37.p13 with BWA-MEM ^94,95^. Single nucleotide variants (SNV) and insertion/deletion (INDEL) sequence variants were identified with the Genome Analysis Toolkit ^96^. The dataset was then lifted-over to hg38 for ease of use.

WES data for 1473 subjects was received in GDS format and converted to variant call format (VCF) using seqGDS2VCF ^97^. Variants were functionally annotated with SnpEff version 5.2 (build 2023-09-29 06:17) ^98^ using the GRCh38.99 reference genome. Annotated VCFs were indexed with tabix and converted to PLINK 1.9’s binary files (.bed/.bim/.fam) to enable genome-wide association analysis. Multiallelic variants were normalized using bcftools v1.11, which split multiallelic sites into multiple biallelic records. All downstream analyses used this normalized dataset, applying additional variant-level filters appropriate to each analysis stage. As of June 2026, 256 subjects with WES data had been classified with a clinical cognitive superager status by RADCO. 1292 subjects from the AFAR dataset had a secondary phenotype available (cognitively normal vs. cognitively impaired).

To identify cryptic relatedness and potential familial clusters, pairwise kinship coefficients were estimated using PLINK 2.0’s --make-king-table function, which implements the KING-robust method ^99^. First-degree relationships (parent-child or full siblings) were identified by kinship coefficients above 0.177 for which there were 46 relational pairs and second-degree relation was defined as a kinship coefficient between 0.125 to 0.177 for which there were zero in our cohort. Cutoffs were set as per the Plink2 documentation ^99^. Five first-degree family groups were detected, defined as clusters where all individuals shared kinship coefficients above 0.177. Allele frequencies were subsequently summarized within each family group and considered during the analyses to assess whether shared rare alleles arose from familial inheritance rather than independent occurrence related to the target endophenotype; a full summary of this family analysis can be found in Supplementary Table B2. For the single-variant and burden analyses, principal components were computed from an LD-pruned variant set to control for population structure; the number of principal components retained as covariates was determined by the scree plot of computed eigenvalues.

To place RADCO whole-exome samples in a common ancestry framework, we projected study genotypes onto the principal-component (PC) space of the 1000 Genomes Project Phase 3 reference panel ^100^. Reference and study variants were harmonized on hg38 and intersected by chromosome and position. The reference panel was restricted to the lifted 1000 Genomes common-variant set used for ancestry inference. Cryptically related reference individuals were removed using KING-robust kinship estimates ^99^, and reference outliers were excluded after an initial autosomal singular value decomposition (SVD), using the local outlier statistic S computed with bigutilsr::prob_dist on the reference PC scores, retaining individuals with S < 0.5. Study samples were then projected into the reference PC space using optimal ancestry projection (OADP) ^14^ as implemented in bigsnpr (bed_projectPCA) ^101^, which matches variants between reference and target data, recomputes the reference SVD on the matched loci, and projects study genotypes without refitting the reference basis on study samples. Projected PCs were visualized with reference samples colored by 1000 Genomes super-population (AFR, AMR, EAS, EUR, SAS) and RADCO participants colored by self-reported race from study metadata. This ancestral projection, Supplementary Figure 1, in 245 subjects retaining their European labels, 5 subjects being re-labeled as European instead of their self-reported races and 6 subjects retaining their labels as non-European. This projection was used to assess overall genetic ancestry and population structure relative to global reference populations; it was complementary to, and separate from, the LD-pruned principal components generated within RADCO and included as covariates in association analyses.

### Single Variant Association Analysis

Single-variant association testing was conducted to identify variants associated with cognitive superager status using a case–control design. Sample-level QC removed one individual that exceeded our heterozygosity and missingness thresholds, leaving 249 samples for variant-level processing. To control for population structure and possible batch effects, we calculated 20 principal components in PLINK v2.0.0-a.6.9LM from 83,046 LD-pruned, autosomal, biallelic SNPs identified by independent-pair pruning with a 50-SNP window, 5-SNP step, and an LD threshold of 0.5 (input set: variants with genotype call rate ≥ 98%, HWEP ≥ 1 × 10^−6^, and minor allele frequency ≥ 1%). Variant filtering for the single-variant analysis applied the following criteria, in order: (i) Hardy–Weinberg equilibrium (HWE)P ≥ 1 × 10^−10^ assessed in the 114 average-cognition control samples (2,061 variants removed), (ii) genotype call rate ≥ 98% (≤ 2% missingness; 116,094 variants removed), and (iii) minor allele frequency ≥ 0.01% to retain high-confidence rare variants and remove monogenic variants (corresponds to a minor allele count of 5, 855,144 variants removed). After variant- and sample-level QC, 418,278 variants remained in 249 samples.

Restricting to participants with complete covariate data left a final analysis set of 231 individuals (129 cognitive superagers, 102 average-cognition controls). Logistic regression was performed using REGENIE v3.5 ^43^ with cognitive superager status as the binary outcome and gender, cognitive age (treated as the midpoint of deidentified age categories; e.g., 100–104 → 102), and the first three principal components (PC1–PC3, retained based on inspection of the scree plot of eigenvalues) as covariates. Analyses were run with a block size of 1,000 and approximate Firth correction enabled at a P-value threshold of 0.05 to mitigate small-sample bias.

### Gene-based Burden Analysis

Rare genetic variants individually tend to lack the statistical power to reach exome-wide significance; to account for this, gene-based burden testing was performed. We employed REGENIE v3.5 ^43^ to aggregate rare variant effects within genes for cognitive superager status. Variant-level QC for the burden analysis was deliberately less stringent than for the single-variant analysis to preserve sparse rare-variant signal: (i) HWEP ≥ 1 × 10^−15^ (mid-p adjustment) assessed in the 114 average-cognition control samples (1,149 variants removed); (ii) genotype call rate ≥ 90% (≤ 10% missingness; 48,160 variants removed); and (iii) minor allele frequency ≥ 0.01% to remove monogenic variants (882,563 variants removed). After QC, 459,705 variants remained, aggregated into 29,376 gene sets across the same 231-individual analysis cohort used in the single-variant analysis (129 cognitive superager cases, 102 average-cognition controls).

To further distill our analysis to variants most likely to have the largest effect, variant functional categories were assigned based on SnpEff annotation. Loss-of-function (LoF) variants were defined as those with SnpEff annotations including frameshift_variant, splice_acceptor_variant, splice_donor_variant, splice_region_variant, start_lost, stop_gained, or stop_lost. Missense variants included missense_variant and missense_variant&splice_region_variant annotations, and all other functional annotations were coded as “other.” We restricted our rare-variant analysis to predicted LoF and missense variants, as these mutation classes are most consistently associated with human disease phenotypes and most likely to exert meaningful biological effects ^102–104^. Synonymous variants were placed in the “other” category as they are functionally silent at the protein level and serve as neutral controls in variant-class enrichment analyses ^105^.

A gene-based set list was generated by grouping annotated variants by gene symbol using REGENIE’s gene-set specification format. Four masks were manually defined in Supplementary Table B9 from the functional categories above: (i) loss-of-function only, (ii) missense only, (iii) combined loss-of-function + missense, and (iv) “other” (synonymous and non-coding). Variants were stratified by allele frequency into bins of MAF ≤ 0.5%, ≤ 1%, ≤ 5%, and singletons, with rare variants of MAC ≤ 10 collapsed within masks. Variant-level weights were assigned using a Beta(MAF; 1, 25) function to upweight rare variants. The final regression model used the binary cognitive superager status as the endophenotype, with gender, age, and PC1–PC3 as covariates and approximate Firth correction applied to account for small-sample bias; a block size of 1,000 was used and a minimum MAC threshold of 5 was applied as in the single-variant analysis.

REGENIE’s RGC-GENE-P test derives a single gene-level omnibus P-value by combining results from multiple burden tests, including ACATV (collapsing burden) and SKATO-ACAT (additive and heterogeneous rare-variant effects), via the Adaptive Cauchy Combination Test (ACAT). By testing across multiple frequency-stratified masks and applying joint ACAT combination, this framework maximizes statistical power to identify genes in which aggregated variants, regardless of frequency or functional impact, are associated with the cognitive superager endophenotype, while controlling Type I error through Firth correction for small, expected cell counts ^43^. REGENIE also reported that 28,786 catalogued sets included variants absent from the exome panel and/or not mappable to the annotation file, and that a subset of 2,209 sets contained only “unknown” variants under the mask rules and were ignored, issues that mainly affect which genes can accumulate detectable signal rather than the validity of P-values for successfully tested sets ^43,86^. Of the 29,376 gene sets tested, 6,765 (23%) yielded no gene-level p-value, predominantly because the constituent variants either failed to match the genotype/annotation data (93% of dropped sets) or, for genes with only 1–2 rare variants, failed to meet the minimum mask allele count (MAC ≥ 5) required for burden mask construction.

To assess whether established Alzheimer’s disease risk and protective signals showed consistent effects in the RADCO cognitive superager cohort independent of exome-wide significance, we performed a targeted lookup of known AD-associated variants and genes against both the single-variant and gene-based burden summary statistics described above. The lookup set comprised the 109 variants reported by Bellenguez et al. ^10^, together with the two *APOE* variants rs429358 and rs7412, which jointly define the ε2/ε3/ε4 alleles ^106^. Each Bellenguez index variant was additionally mapped to a single gene for the burden-test comparison using the prioritized causal gene reported by Bellenguez et al. ^10^; where multiple genes were reported per locus, the highest-confidence assignment was used. For the single-variant comparison, each lookup variant was matched to our REGENIE additive single-variant statistics by chromosome and position. Effect alleles were harmonized so that the tested allele in our statistics matched the allele reported by Bellenguez et al. ^10^, with β estimates and standard errors sign-flipped where necessary. Under a protective-cognitive-superager hypothesis, AD-risk alleles are expected to be depleted in cognitive superagers relative to controls, yielding negative β values in our case-versus-control comparison.

For the known AD gene-based comparison, each Bellenguez-prioritized gene was matched against our REGENIE GENE_P statistics. Because GENE_P is a two-sided aggregate statistic that does not return a directional effect estimate, direction-of-effect concordance was not assessed at the gene level; the gene-based comparison was restricted to identifying overlap between Bellenguez-prioritized genes and genes nominally associated in our burden analysis.

### Rare Variant Allele Frequency Enrichment Testing

Allele frequency analyses were performed using the normalized RADCO WES data in which all multiallelic sites had been decomposed into biallelic variants using bcftools as previously described. Allele frequencies were then calculated across the entire cohort as well as within cognitive superagers and average-cognition subjects using PLINK 2.0’s --freq command, yielding subgroup-specific allele frequency (AF) and allele observation numbers (AN). Variant-level missingness was calculated for the cohort as a whole to identify sites with poor call rates. Variants with cohort-wide call rate < 90% would have been excluded prior to the enrichment test had any met that threshold. For each variant, subgroup-specific allele frequencies and alleles observed were compared across RADCO cognitive superagers, RADCO average-cognition subjects, and external reference data from gnomAD’s non-Finnish Europeans (NFE), allele counts (AC) were approximated as AN x AF, rounded to the nearest integer to represent counts of alternative alleles. 6 subjects including 2 cognitive superagers whose ancestry did not fall under gnomAD’s NFE definition were excluded from the allele frequency enrichment analysis.

The Genome Aggregation Database, or gnomAD, v4.1 is an updated reference population released in April of 2024, aggregating and harmonizing exome sequencing data from over 730,000 participants including 416,555 UK Biobank and 314,392 non-UK biobank exomes. The v4.1 exome VCF files and their accompanying index files were downloaded directly from the gnomAD Browser provided by the Broad Institute ^107^. These files contain genotypes and allele frequency information across the cohort enabling comparison of variant frequencies within the RADCO group and this large reference population. A subset of the entire gnomAD reference that focuses on the Non-Finnish European (NFE) population was used as it aligns most closely with the demographics of the RADCO cohort. Finnish people are excluded from this mixture due to the prevalence of Finnish Disease Heritage diseases ^108^.

Fisher’s exact tests were then applied to 2 × 2 contingency tables of allele counts to test the null hypothesis of there being no difference in allele frequency between groups, consistent with standard practice in rare-variant case-control analyses where expected cell counts are small and asymptotic approximations are unreliable ^85,86^. Three pairwise Fisher’s exact comparisons were conducted in this manner: (i) RADCO cognitive superagers vs. gnomAD NFE, (ii) RADCO average-cognition vs. gnomAD NFE and (iii) RADCO cognitive superagers vs. RADCO average-cognition. The primary aim was to identify variants for which cognitive superagers showed a statistically significant allele frequency difference relative to gnomAD, while the average-cognition group did not differ significantly from gnomAD, consistent with enrichment specific to the cognitive superager endophenotype rather than general cohort batch effects. Resulting Fisher’s exact P-values were then Benjamini-Hochberg corrected for multiple comparisons.

Analyses started from 1,224,073 exonic variants. Variants that were not screened across all three groups were removed because their rarity and frequency could not be properly benchmarked between groups. In addition, common variants were excluded by filtering out variants with gnomAD AF_NFE_ ≥ 0.001 or gnomAD AF_NFE_ ≤ 0.999, restricting the analysis to the rarest exonic variants in the reference population. Variants were retained only if they demonstrated a statistically significant difference in allele frequency via Fisher’s exact test between the RADCO cognitive superager group and the gnomAD NFE reference population (Benjamini–Hochberg adjusted Fisher’s exact P < 0.01) while simultaneously showing no significant difference between the RADCO average-cognition group and gnomAD NFE (Benjamini–Hochberg adjusted Fisher’s exact P > 0.01). This criterion was designed to isolate variants whose frequency enrichment is specific to the cognitive superager phenotype rather than batch effects.

Further, we filtered out variants with a minor allele count less than 4 within the cognitive superager cohort resulting in 976 candidate variants. To test whether the functional-consequence composition of these candidate variants deviated from expectation, we compared them to a frequency-matched empirical background comprising all 870,018 rare exonic variants retained after the gnomAD NFE rarity filter (AF_NFE_ < 0.001) but prior to superager allele-frequency pruning. Each variant was assigned a SnpEff consequence class, and the missense-to-synonymous ratio of the candidate coding subset was compared to that of the background using a two-sided Fisher’s exact test. This matched design controls for the whole-exome capture footprint, the rarity threshold, and the annotation pipeline, isolating any functional-class enrichment attributable to the superager phenotype.

Candidate SNPs were further filtered to retain only variants predicted to alter protein structure or function. Using SnpEff annotations, we retained variants in the missense, protein-truncating (stop_gained, stop_lost, start_lost, frameshift_variant), and splice-disrupting (splice_acceptor_variant, splice_donor_variant, splice_region_variant in combination with a synonymous, intronic, or missense context) effect classes, mirroring the high- and moderate-impact tiers used in standard rare-variant case-control analyses ^102,104^. Variants failing HWE at a threshold of 1 x 10^-6^ within the entire RADCO WES cohort (n = 1473) were removed as well as those variants for which all alternative allele carriers came from the same first-degree family group. This filtering resulted in 114 variants that were then split into confidence tiers 1 & 2 by the cognitive superager vs. gnomAD NFE comparison at a threshold of P_adj_ = 1.0 x 10^-8^.

Five candidate variants from the allele-frequency analysis, two tier 1 and three tier 2, mapped to the HLA region leading to further investigation. Reliable HLA imputation requires chromosome 6 whole-genome sequence data that was not widely available in the cognitive superager allele frequency cohort but was available for a broader AFAR subset. We performed an accessory HLA association analysis using the secondary endophenotype of cognitively normal (n = 524) versus cognitively impaired (n = 248) status (n = 772 with covariate-complete data), 280 of which were centenarians (112 cognitively impaired, 168 cognitively normal). Classical two-field HLA types at seven loci were imputed from chromosome 6 WGS genotypes using HIBAG v1.44.0 (European reference model) ^109,110^, with population structure adjusted using WES-derived PC1–PC6 from the full RADCO WES panel (n = 1,473). For each locus, the lead allele was tested in logistic regression (additive dosage; sex, age, PCs) and Benjamini–Hochberg FDR was applied across seven lead-allele tests.

To evaluate whether allele frequency candidate variants are predicted to alter gene regulation or splicing in neural contexts, we performed in silico regulatory profiling of all 114 candidates using AlphaGenome ^111^ in brain (UBERON:0000955) and neuron (CL:0000540) ontologies. For each variant, we computed the maximum absolute predicted difference between reference and alternate alleles across RNA-seq, CAGE, splice-site, DNase, and histone-modification tracks within a ± 262 kb window. Splice-site scores are probabilities (0–1); the remaining modalities are normalized predicted-coverage signals in assay-specific, relative units and are therefore not directly comparable across columns. This analysis was intended to support functional interpretation of AF candidates rather than to provide independent validation of cognitive superager enrichment, because all 114 variants had already been ascertained on allele-frequency grounds. For each modality, we retained the maximum absolute difference between reference and alternate allele predictions across the two cell ontologies; larger scores indicate that the model predicts a greater regulatory or splicing perturbation for the alternate allele relative to reference. Variants were annotated with Ensembl VEP^112^. AlphaGenome scores were merged with cognitive superager carrier frequencies from the European-ancestry subset with non-missing cognitive superager phenotype (n = 138 cognitive superagers, n = 118 average-cognition controls).

To explore whether the prioritized candidate genes converged on shared biological processes, we performed over-representation analysis (ORA) on the unique protein-coding genes carrying the 114 candidate variants using Enrichr ^113,114^ via the gseapy Python interface^115^. Gene symbols were queried against four annotation libraries, Gene Ontology Biological Process ^116^, KEGG ^117^, Reactome ^118^, and MSigDB Hallmark ^119^, and enrichment was assessed by Fisher’s exact test with Benjamini–Hochberg FDR correction applied within each library, with terms at FDR < 0.05 considered significant.

### Computational Setup

Analyses were performed on the ERISTwo high-performance computing cluster (Mass General Brigham Enterprise Research Infrastructure & Services), a CentOS 7 Linux cluster with approximately 75 compute nodes. Job submission was managed via Slurm. Computational workflows were executed in a Conda (v25.7.0) virtual environment running Python v3.11.9, R version v4.4.0, and GNU bash v4.2.46. Key package versions are provided in the environment specification supplementary file B10.

## Supporting information

Supplementary Figues

## Data Availability

gnomAD v4.1 exonic and 1000 Genomes reference data is publicly available. Study Data will be available from the ELITE portal.

https://gnomad.broadinstitute.org/downloads

https://www.internationalgenome.org/data/

https://eliteportal.synapse.org/

## Funding and acknowledgements

Data, methods used, and/or research reported in this publication were provided in whole or in part by the Resilience/Resistance to Alzheimer’s Disease in Centenarians and Offspring (RADCO) Study. This project was supported by NIH Research Grants U19AG073172, U19AG023122, and UH2/UH3AG064704 funded by the National Institute on Aging (NIA), and the McKnight Brain Research Foundation (MBRF), the American Federation for Aging Research, and the Cure Alzheimer’s Fund.

The investigators within RADCO contributed to the design and implementation of the study and/or provided data but may not have directly contributed to the analysis or writing of this publication. A complete listing of RADCO key investigators can be found at: https://www.bumc.bu.edu/centenarian/files/2025/12/RADCOAcknowledgement-Policy-for-Publications.pdf. The authors used Github Copilot to assist with analysis scripts and language editing during manuscript preparation, and for literature searches. All AI-assisted output were reviewed and verified by the authors. The content of this publication is solely the responsibility of the author(s) and does not necessarily represent the official views of the National Institutes of Health.

## Conflicts of Interest

The authors declare that they do not have any conflicts of interest with the content of this article.

## Data Availability

Study Data will be available from the ELITE portal. gnomAD v4.1 exonic reference data is publicly available from https://gnomAD.broadinstitute.org/downloads.

## Ethics Statement

Participants were enrolled in the New England Centenarian Study (NECS), Longevity Consortium Centenarian Project (LCCP), Integrative Longevity Omics (ILO), and/or the Resilience/Resistance to Alzheimer’s Disease in Centenarians and Offspring Study (RADCO). The NECS and LCCP studies were approved by the institutional review board at Boston University Medical Campus. The ILO and RADCO studies were approved by the single institutional review board at Albert Einstein College of Medicine. All participants or their legally authorized representatives provided written informed consent.

## Notes

### Competing Interest Statement

The authors have declared no competing interest.

