## Supplementary Figues for "Enrichment of Rare Exonic Variants in Cognitive Superagers from Centenarian Families"

Supplementary Figure 1:

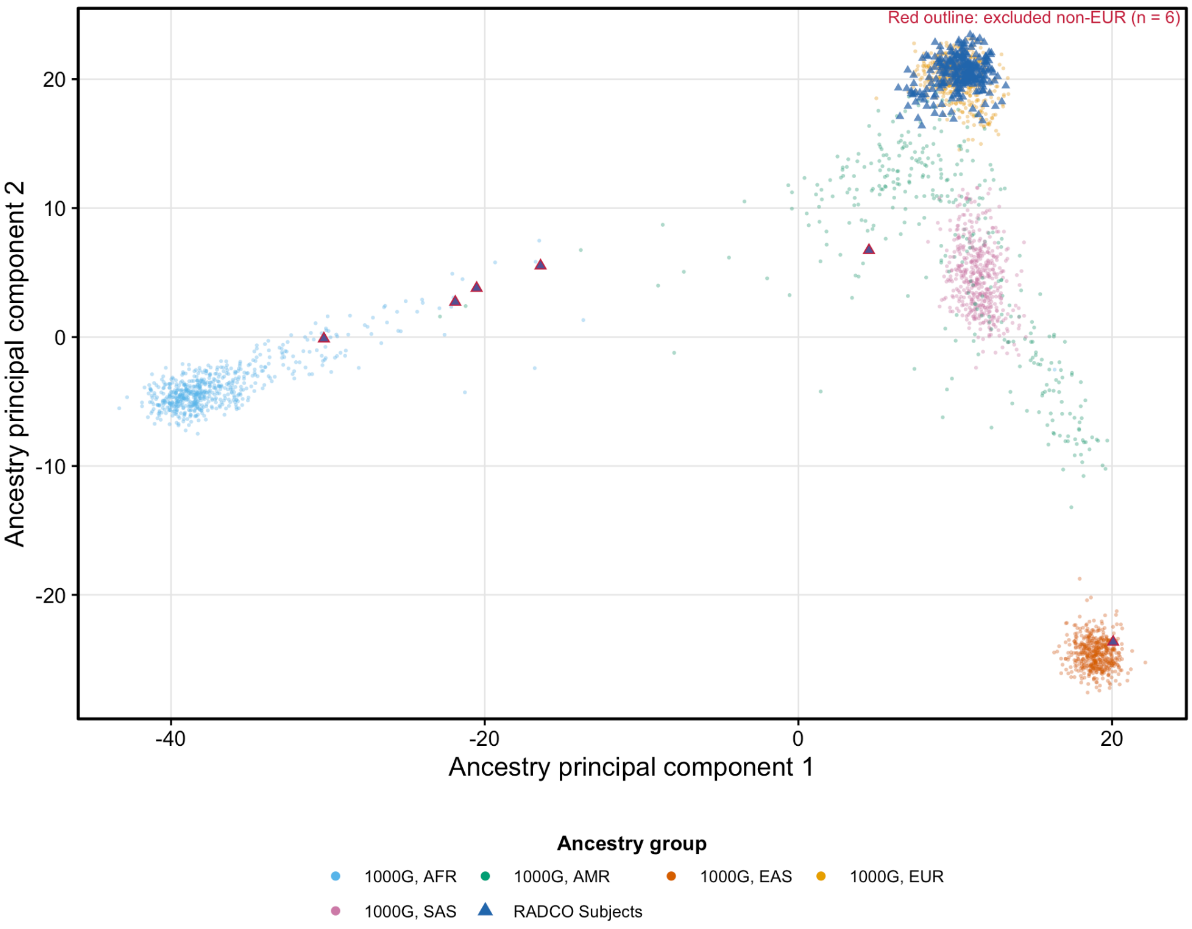


**Supplementary Figure 1: Genetic ancestry projection of RADCO Superager whole-exome sequencing (WES) samples onto the 1000 Genomes Project Phase 3 reference (hg38).** Principal components were estimated with optimal ancestry projection (OADP; bigsnpr) using a lifted 1000 Genomes common-variant reference (n = 2425 after relatedness filtering and outlier removal). RADCO study samples (n = 256) were projected into the reference PC space using 46,029 SNPs intersected on chromosome:position with the reference panel, followed by standard QC. Reference individuals (circles) are colored by 1000 Genomes super-population (AFR, African; AMR, Admixed American; EAS, East Asian; EUR, European; SAS, South Asian). RADCO participants are represented by triangles, with the non-European subjects being highlighted with red as they were excluded from the primary analysis.

AFR, African; AMR, Admixed American; EAS, East Asian; EUR, European; OADP, optimal ancestry projection; PC, principal component; SAS, South Asian; WES, whole-exome sequencing.
